# OMOP CDM for breast cancer research: transforming the Breast Cancer Now Biobank data

**DOI:** 10.64898/2025.12.14.25342223

**Authors:** Maryam Abdollahyan, BCNB-BCI, Claude Chelala

**Affiliations:** Centre for Cancer Biomarkers and Biotherapeutics, Barts Cancer Institute, Queen Mary University of London, London EC1M 6BQ, United Kingdom; Barts Centre of the Breast Cancer Now Biobank, Barts Cancer Institute, Queen Mary University of London, London EC1M 6BQ, United Kingdom

## Abstract

Common data models (CDMs) are essential for health data standardisation, which facilitates the governance and management of data, improves data quality and enhances the findability, accessibility, interoperability and reusability of data. They allow researchers to efficiently integrate health datasets and perform joint analysis on them, promoting collaboration and maximising translation of research outputs for patients’ benefit. We describe the process of transforming the biobank data for over 2,850 donors recruited at the Barts Cancer Institute (BCI) site of the Breast Cancer Now Biobank (BCNB) - the UK’s first national breast cancer biobank hosting longitudinal biospecimens and associated clinical, genomic and imaging data - into the Observational Medical Outcomes Partnership (OMOP) CDM. Our transformation pipeline achieved high coverage, with 83% of source concepts mapped, and our OMOP CDM achieved a total pass rate of 100% in quality assessments. We present the breast cancer characteristics of the resultant patient cohort. We report several challenges faced during the transformation process and explain how we addressed them, and discuss the strengths and limitations of adopting the OMOP CDM for breast cancer research. The OMOP-mapped BCNB-BCI dataset is a valuable resource that can now be explored and analysed alongside other health datasets.

## Introduction

Standardisation facilitates the governance and management of health data, and improves data quality in terms of consistency and accuracy. It also ensures compliance with the FAIR principles of findability, accessibility, interoperability and reusability, which maximises translation of research outputs. Standardisation allows data from multiple sources to be linked more efficiently, enabling researchers to seamlessly integrate diverse datasets and perform joint analysis on them. This promotes collaboration among various stakeholders, accelerating discoveries that ultimately enhance patient care and outcomes.

Central to semantic interoperability are data standards (e.g., Fast Healthcare Interoperability Resources (FHIR)(1)), common data models (CDMs) (e.g., Observational Medical Outcomes Partnership (OMOP)(2)) and biomedical ontologies (e.g., Systematized Nomenclature of Medicine Clinical Terms (SNOMED CT)(3)). The OMOP CDM - widely adopted by health data initiatives - aims to harmonise observational health datasets. Transforming an existing health dataset into the OMOP CDM involves standardising both its structure and content. Records are mapped to concepts from the Observational Health Data Sciences and Informatics (OHDSI) standard vocabularies. Majority of these vocabularies are external ontologies commonly used in healthcare (e.g., SNOMED CT), while the rest are internal OMOP ontologies. Mapped records are hosted in a person-centric relational database that comprises of tables containing information which belongs to domains typically found in observational health data (conditions, drug exposures, procedures, measurements etc.). This uniform representation of data allows for the shared analysis of disparate observational health datasets.

Here, we present the transformation of the Breast Cancer Now Biobank (BCNB) - the UK’s first national breast cancer biobank with multiple recruitment sites in England, Wales and Scotland - data into the OMOP CDM. BCNB hosts longitudinal biospecimens and associated clinical, genomic and imaging data from breast cancer patients at different stages of their care pathway (4). We describe the process of transforming the BCNB-BCI dataset - data for donors recruited at the Barts Cancer Institute (BCI) site of BCNB - into the OMOP CDM, and discuss the challenges faced and opportunities for improvement.

## Methods

### Data source

To date, the BCI site of BCNB has recruited over 3,890 donors, including patients with breast cancer or benign breast conditions and individuals at high risk of breast cancer. The Biobank routinely collects longitudinal biospecimens and associated data from its donors. Biospecimens stored by the Biobank include liquid samples (e.g., plasma, serum, buffy coat, urine and faeces), tissue samples (e.g., fresh frozen tissue, FFPE blocks and H&E-stained slides) and cells. Clinical data manually curated by the Biobank include demographics, medical history and lifestyle, diagnoses, procedures and treatments. Biospecimens from a subset of the donors have been sequenced (germline and somatic whole genome sequencing or RNA-Seq). In addition, for many of the donors, imaging data (e.g., radiology and H&E-stained whole slide images) are available.

Data hosted by BCNB-BCI are linked to electronic health records (EHRs) from Barts Health NHS Trust (BH) - one of the largest NHS trusts in London with five hospitals serving over 2.5 million people across East London. EHRs from BH include demographics, visits, diagnoses, procedures, electronic prescriptions and free text data such as imaging and pathology reports. The BCNB-BCI data are also linked to GP records from the East London NHS Discovery programme as well as sequencing data from the 100,000 Genomes Project (100K GP) (5). Majority of these linked external data are coded using an OHDSI standard vocabulary, which guided our choice of target vocabularies.

### Extract, transform and load (ETL) process

The first step in our ETL process was extraction of the source data hosted in the BCNB Research Electronic Data Capture (REDCap) (6) system. Data for each record (donor) are captured in multiple instruments (forms or tables) which are grouped together under an event (clinical endpoints of disease progression and response: primary breast cancer, local and regional recurrence, metastasis and remission). In this project, we transformed only the BCNB-BCI clinical data, excluding instruments that contain information on the Biobank’s biospecimens. Two instruments were partly mapped to OHDSI standard vocabularies since their fields are mostly operational (e.g., donor consent, audit history) or calculated, i.e., their value is derived from other fields (e.g., time from primary breast cancer diagnosis to metastasis). Donors who have withdrawn their consent, newly recruited donors for whom no clinical data have been entered yet, and donors with only archival biospecimens (e.g., archival FFPE blocks) and limited clinical data were excluded from the ETL process.

The next step in our ETL process was mapping the source data to OHDSI standard vocabularies. Fields in each data collection instrument can be considered as belonging to one of two main categories: general and breast cancer-specific. Neither category used an ontology to represent its values prior to mapping.

Value of general fields (e.g., date of birth) is available for all the Biobank’s donors, regardless of their disease status. These fields were mainly mapped to SNOMED CT concepts. In addition to being an OHDSI standard vocabulary, the majority of EHRs linked to the Biobank’s data are coded using SNOMED CT, which allows us to compare the Biobank’s manually curated data and linked external data. Some general fields were mapped to a different vocabulary (that is, directly to one of the internal OMOP ontologies) if their corresponding SNOMED CT concept had become invalid (e.g., remapped to another concept or deprecated) or non-standard, or belongs to the wrong domain.

Values of breast cancer-specific fields are primarily derived through manual curation from free text data such as clinical reports and letters. The Biobank’s data curators extract this information from EHRs and enter them as structured data in the Biobank’s database. Similar to general fields, where possible, we mapped these fields to SNOMED CT concepts, and if a SNOMED CT concept could not be found, we mapped them to a cancer-specific vocabulary. For example, tumour stage was mapped to the Cancer Modifier vocabulary, and chemotherapy regimens were mapped to the HemOnc vocabulary. Other standard vocabularies used in the mapping process are Logical Observation Identifiers Names and Codes (LOINC) (10) and Unified Code for Units of Measure (UCUM) (11).

All mappings between source codes and OMOP standard concepts were performed manually using the Athena (7) database - a repository of OMOP CDM vocabularies maintained by the OHDSI team - and custom Python scripts, prioritising SNOMED CT concepts that are frequently used in the linked EHRs. We documented the mappings using the WhiteRabbit (8) and Rabbit-In-a-Hat (9) applications - tools developed by the OHDSI team for generating ETL specification documents. Table 1 shows an overview of the information available in each data collection instrument and the ontologies chosen to represent values of its fields.

**Table 1.** Overview of the information in each data collection instrument and the ontologies representing it.

| Data Collection Instrument | Source Contents | Target Vocabulary |
| --- | --- | --- |
| Identification | Donor details including identifiers, DOB, biological sex, ethnicity and survival status. | SNOMED CT |
| Demographics |  |  |
| Survival Status |  |  |
| Family Medical History | Family history of cancer, especially breast and ovarian cancers. | SNOMED CT |
| Medical History | Current and past (non-breast cancer) conditions, procedures and medications.<br>Menopausal status and HRT.<br>BMI.<br>History of smoking, alcohol consumption and drug use.<br>Parity and history of breastfeeding and contraception use. | SNOMED CT<br>LOINC<br>UCUM |
| Lifestyle |  |  |
| Presentation | Breast cancer presentation (detection and symptoms) and diagnosis. | SNOMED CT |
| Diagnosis |  |  |
| Biopsy Procedure | Breast cancer biopsy and surgery procedure details.<br>Histopathology report including invasive, in-situ and benign features, and lymph node and receptor status.<br>Breast cancer imaging procedure details and report including BI-RADS score. | SNOMED CT<br>LOINC<br>UCUM |
| Surgery Procedure |  |  |
| Imaging Procedure |  |  |
| Diagnostic Summary | Breast cancer tumour size, grade and TNM stage. | SNOMED CT<br>Cancer Modifier |
| Therapy | Breast cancer chemotherapy, radiotherapy, biological and endocrine therapies and palliative care details.<br>Treatment response and side effects. | SNOMED CT<br>HemOnc |
| Follow-up | Last consultation details and report. | SNOMED CT<br>LOINC |

Finally, output of the mapping process was imported into a PostgreSQL database. We used an R package (12) developed by the OHDSI team to create the OMOP CDM tables (that is, to generate the Data Definition Language (DDL) scripts) for this database.

### Validation

We visualised and evaluated the results of the ETL process using the ACHILLES (13) and Data Quality Dashboard (DQD) (14) R packages - tools developed by the OHDSI team for characterising and assessing the data quality of an OMOP CDM instance. DQD checks the data for plausibility, conformance and completeness, and computes a pass percentage for each category. All the issues found by DQD were investigated and resolved, where applicable.

Figure 1 shows an overview of the transformation process.

**Figure 1.**
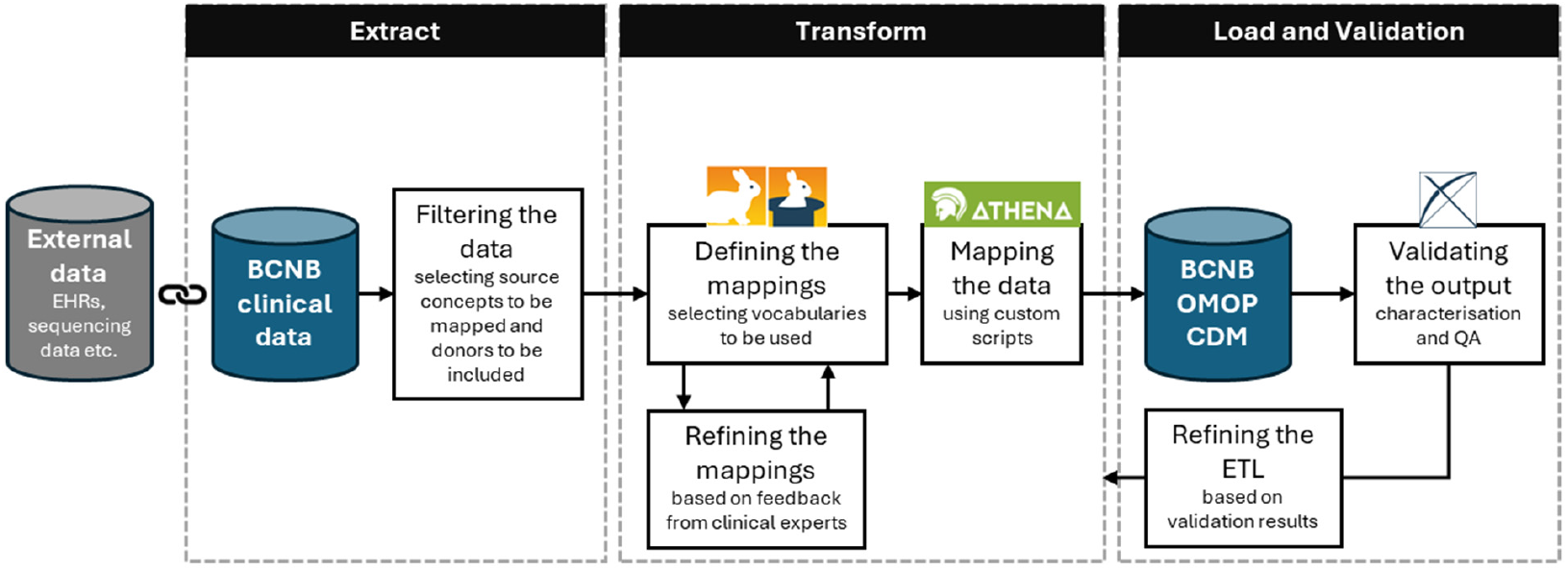
Overview of the transformation process

## Results

We identified 2,923 BCNB donors who met the inclusion criteria outlined in the Methods section, and transformed the clinical data for 2,871 donors into the OMOP CDM. We excluded 52 donors of unknown ethnicity since the *race_concept_id* field in the *person* table is a mandatory field.

### Transformed BCNB-BCI cohort

We characterised the resultant cohort through statistics of demographics and family history, primary and subsequent diagnoses, and tumour characteristics. Table 2 shows the breast cancer characteristics of the transformed BCNB-BCI cohort (release date: October 2025). The median age at diagnosis of primary breast cancer for patients (or age at recruitment for other donors) was 56 years, with a median follow-up time of 6 years. The BCNB-BCI cohort was predominantly female (99.3%) and White (59%), with 14.1% Asian or Asian British and 12.1% Black or Black British donors. Majority of the donors were diagnosed with breast cancer (93.8%), while smaller proportions had benign, high-risk or normal diagnoses. Among donors with recorded tumour characteristics, grade II tumours were the most common (39%), and ER-positive and HER2-negative tumours accounted for 64.6% and 59.6%, respectively.

**Table 2.** Breast cancer characteristics of the transformed cohort. The ‘Other’ category includes cases such as donors who underwent cosmetic breast surgery.

| Characteristic | Value for Total Donors | Percentage of Total Donors |
| --- | --- | --- |
| **Age (years) [median]** | 56 | - |
| **Biological Sex** | - |  |
| Female |  | 99.30 |
| Male |  | 0.70 |
| **Ethnicity** | - |  |
| Asian or Asian British |  | 14.11 |
| Black or Black British |  | 12.09 |
| Mixed |  | 2.41 |
| White |  | 59.02 |
| Other |  | 5.54 |
| Not Stated |  | 6.83 |
| **Family History of Breast Cancer** | - |  |
| Yes |  | 17.32 |
| No |  | 15.33 |
| Not Stated |  | 67.35 |
| **Diagnosis** | - |  |
| Primary |  | 93.76 |
| Benign |  | 1.74 |
| High Risk |  | 2.65 |
| Normal Control |  | 0.7 |
| Other |  | 1.15 |
| **Subsequent Event** | - |  |
| New Primary |  | 3.45 |
| Recurrence |  | 8.5 |
| Metastasis |  | 8.71 |
| None |  | 73.21 |
| Not Applicable |  | 6.24 |
| **Grade** | - |  |
| I |  | 12.44 |
| II |  | 39.02 |
| III |  | 21.99 |
| Not Stated |  | 20.31 |
| Not Applicable |  | 6.24 |
| **Nodal Status** | - |  |
| Positive |  | 53.21 |
| Negative |  | 28.26 |
| Not Stated |  | 12.29 |
| Not Applicable |  | 6.24 |
| **Receptor Status** | - |  |
| **ER** |  |  |
| Positive (scores 4-8) |  | 64.56 |
| Borderline (score 3) |  | 0.94 |
| Negative (scores 0-2) |  | 11.67 |
| Not Stated |  | 16.59 |
| Not Applicable |  | 6.24 |
| **HER2** |  |  |
| Positive (score 3+) |  | 6.86 |
| Borderline (score 2+) |  | 9.41 |
| Negative (scores 0-1+) |  | 59.65 |
| Not Stated |  | 17.84 |
| Not Applicable |  | 6.24 |
| **Follow-up Time (years) [median]** | 6 | - |

We also created subgroups of interest and characterised them through, for instance, the sequence of treatments received. Figure 2 shows the treatment pathway (up to third line) for a subgroup of 104 patients from the transformed BCNB-BCI cohort with recorded treatment information after being diagnosed with metastatic breast cancer. Majority of the patients received chemotherapy or biological therapy followed by endocrine therapy.

**Figure 2.**
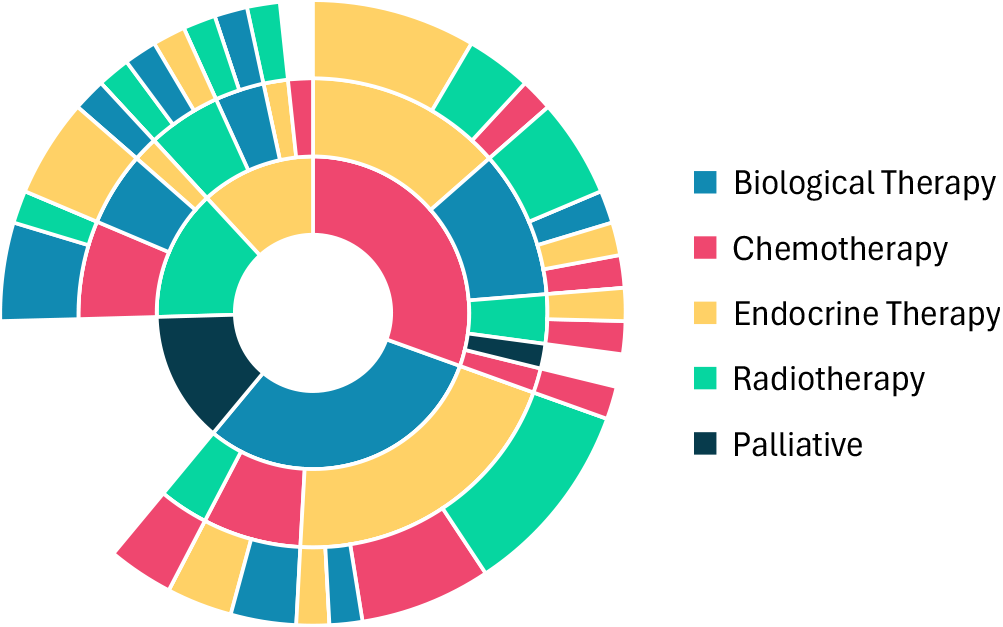
Treatment pathway after metastatic breast cancer diagnosis for a subgroup of patients from the transformed cohort

### Mapping metrics

We mapped 470 out of 562 (83.6%) source concepts. Table 3 shows the proportion of mapped concepts for each data collection instrument. Mapping rates were particularly high for the *Identification, Family Medical History, Presentation, Diagnosis, Imaging Procedure* and *Follow-up* instruments, with each achieving a mapping rate above 95%. Lowest mapping rates were observed for the *Demographics* (40%) and *Medical History* (78.9%) instruments. We discuss the reasons why a match could not be found for the unmapped concepts in the next section. The remaining six instruments achieved a mapping rate between 79% and 94%.

**Table 3.** Proportion of mapped concepts for each data collection instrument.

| Data Collection Instrument | No. of Source Concepts | No. of Mapped Concepts | Mapping Rate (%) |
| --- | --- | --- | --- |
| Identification | 5 | 5 | 100.00 |
| Demographics | 25 | 10 | 40.00 |
| Survival Status | 5 | 4 | 80.00 |
| Family Medical History | 12 | 12 | 100.00 |
| Medical History | 19 | 15 | 78.95 |
| Lifestyle | 32 | 30 | 93.75 |
| Presentation | 10 | 10 | 100.00 |
| Diagnosis | 42 | 40 | 95.24 |
| Biopsy Procedure | 131 | 104 | 79.39 |
| Surgery Procedure | 146 | 118 | 79.73 |
| Imaging Procedure | 25 | 25 | 100.00 |
| Diagnostic Summary | 14 | 13 | 92.86 |
| Therapy | 92 | 80 | 86.96 |
| Follow-up | 4 | 4 | 100.00 |
| **Total** | **562** | **470** | **83.63** |

DQD achieved a pass percentage of 100%, 99% and 99% in the *Plausibility, Conformance* and *Completeness* categories, respectively. Three checks in the *Conformance* category and one check in the *Completeness* category failed. Two of these checks failed since the ‘non-cancer death’ value could not be mapped (that is, the *cause_concept_id* field in the *death* table was set to *0*) for over 5% (maximum percentage of non-compliant records allowed for this check to pass) of the deceased donors. One check failed since 14% of the donors are from an ethnic background that could not be mapped. The remaining check failed since the *ethnicity_concept_id* field in the *person* table was set to *0* for all donors. This field is intended for US-based data only, and therefore, is not applicable here.

Overall, our transformation pipeline produced a high-quality OMOP CDM that accurately represents the BCNB-BCI dataset.

## Discussion

We described the process of transforming the clinical data hosted by BCNB-BCI into the OMOP CDM. Our approach achieved high coverage, with over 83% of fields mapped to OHDSI standard vocabularies. Moreover, our OMOP CDM passed all quality assessment checks with a total pass rate of 100%. However, we faced several challenges during this process. Some of these challenges have also been reported in previous studies (15,16).

### Challenges and limitations

Some fields could not be mapped (or were mapped to *0* indicating no matching concepts) since there exist no accepted concepts to represent them. For example, currently, there are no accepted (valid, standard and belonging to the *Race* domain) concept IDs for the *race_concept_id* field in the *person* table to represent mixed ethnic groups (e.g., the *Mixed White and Black Caribbean* category (17)). Similarly, there is no accepted concept ID for the *ethnicity_concept_id* field in the same table to represent the Ashkenazi Jewish ethnic group - a population with elevated risk of breast cancer due to the high prevalence of BRCA1 and BRCA2 gene mutations. This affected a large proportion of records from BCNB-BCI as a site that recruits donors predominantly from the multi-ethnic population of East London. Ethnicity information is vital for precision medicine and reducing health inequalities.

Values such as ‘not stated’ or ‘patient refused’ are generally not mapped since they are considered the same as ‘unknown’ value. However, it is essential for BCNB that these values are included so cases where the donor chose not to provide information or refused tests or treatment be distinguished from the default unknown cases. Inclusion of these values is also essential when the source data are linked to external data that contain these values since it improves the quality of data linkage.

Although negated values (e.g., absence of a condition) are usually not mapped, we mapped important breast cancer-specific fields that contain such values. These data are frequently requested by applicants to the Biobank when defining cohorts of patients with characteristics of interest. Negated values can be mapped using different approaches, sometimes with no clear guidelines on which approach to follow. For instance, absence of a family history of breast cancer can be represented in few ways, including as a combination of the *observation_concept_id* and *value_as_concept_id* fields in the *observation* table, where *observation_concept_id* is ‘no family history’ (e.g., SNOMED CT code 160266009) and *value_as_concept_id* is ‘breast cancer’ (e.g., SNOMED CT code 254837009), or using only the *observation_concept_id* field, where *observation_concept_id* is ‘no family history of breast cancer’ (e.g., SNOMED CT code 313376005). We followed the OHDSI recommendations, where available, and otherwise, the approach that yielded the best coverage was followed.

Some fields could be mapped to multiple OHDSI standard vocabularies. Without a clear roadmap on how these vocabularies evolve over time (e.g., become deprecated), deciding which one to choose is not trivial. We prioritised vocabularies that are used for coding the EHRs linked to the Biobank’s data, which improves data interoperability. Likewise, some fields could be combined into a single concept. For example, ductal carcinoma in situ (DCIS) and microinvasion can be represented by two concepts (e.g., SNOMED CT code 109889007 and Cancer Modifier code OMOP4999699, respectively) or by a single concept, i.e., DCIS with microinvasion (e.g., SNOMED CT code 443933007). We prioritised concepts that are present in the OMOP-mapped datasets by other data providers to further increase interoperability.

All the vocabularies used for mapping were downloaded from Athena, but they were not necessarily the latest version released by their distributors. Therefore, for instance, any code found via SNOMED CT Browser (18) or related applications (e.g., NHS Digital SNOMED CT Browser (19)) by healthcare professionals who reviewed the mappings that did not exist in Athena as well as any deprecated SNOMED CT code that was still valid in Athena had to be replaced. Similarly, it is possible for these vocabularies to be no longer maintained and their accepted concepts to became invalid or non-standard over time, as is the case with Cancer Modifier and HemOnc. Furthermore, the BCNB data dictionary constantly evolves, for example, to include new breast cancer therapies. Hence, transforming any dataset into the OMOP CDM is not a one-off but a continuous process that requires the source data, OMOP CDM vocabularies and schema, ETL scripts and validation tools to be updated regularly.

### Future work

Precision medicine requires the integration of genomic data into our OMOP CDM. Hence, future work will focus on coding the Biobank’s biospecimen data and genomic variants that are clinically relevant to breast cancer using the OMOP Genomic (20) vocabulary.

## Conclusions

We presented an example of a national cancer biobank’s dataset successfully transformed into the OMOP CDM. The transformation pipeline achieved high coverage and excellent data quality. We reported several challenges faced during the transformation process and how we addressed them. The resulting OMOP-mapped BCNB-BCI dataset becomes a valuable resource for breast cancer research that can be explored and analysed alongside other datasets using emerging methodologies such as federated analysis.

## Data Availability

The data that support the findings of this study are available upon submission of an application to the Breast Cancer Now Biobank.

## Data Availability

The data that support the findings of this study are available upon submission of an application to the Breast Cancer Now Biobank. The data are not publicly available due to ethical and governance restrictions.

## Acknowledgements

The authors would like to thank Barts Health NHS Trust and the East London NHS Discovery programme for their support in the collection of secondary and primary care records. The authors would also like to thank the OPTIMA consortium partners for their valuable feedback and recommendations on the transformation process.

The BCNB-BCI team includes Maryam Abdollahyan, Emanuela Gadaleta, Qianqian Zhu, Sonal Varsani, Catherine McMaster-Christie, Katie Thornton, Naushin Waseem, Anjum Haque, Maritchka Ryniejska, Thamid Yaseen, Iain Goulding, Claude Chelala and Louise J Jones.

## Funding

BCNB is funded by the Breast Cancer Now charity. M.A. is supported by BCNB (grant no. TBYG2E8) and the OPTIMA project (grant no. BTXN1A1R) awarded to C.C.

## Conflicts of Interest

The authors have no conflicts of interest to declare.

## Author Contributions

M.A. extracted the data, implemented the methodology, performed the analysis, generated the tables and figures, and drafted the manuscript. C.C. conceptualised the research question, acquired the funding, supervised the project and edited the manuscript.

## Notes

### Competing Interest Statement

The authors have declared no competing interest.

### Funding Statement

The Breast Cancer Now Biobank (BCNB) is funded by the Breast Cancer Now charity. Maryam Abdollahyan is supported by BCNB (grant no. TBYG2E8) and the OPTIMA (Optimal treatment for patients with solid tumours in Europe through Artificial Intelligence) project (grant no. BTXN1A1R) awarded to Claude Chelala.

### Author Declarations

East of England - Cambridge Central Research Ethics Committee (REC ref. 23/EE/0229) gave ethical approval for this work.

